# Evaluation of three rapid lateral flow antigen detection tests for the diagnosis of SARS-CoV-2 infection

**DOI:** 10.1101/2020.12.30.20249057

**Authors:** AE Jääskeläinen, MJ Ahava, P Jokela, L Szirovicza, S Pohjala, O Vapalahti, M Lappalainen, J Hepojoki, S Kurkela

## Abstract

**Introduction:** The COVID-19 pandemic has led to high demand of diagnostic tools. Rapid antigen detection tests have been developed and many have received regulatory acceptance such as CE IVD or FDA markings. Their performance needs to be carefully assessed.

**Materials and Methods:** 158 positive and 40 negative retrospective samples collected in saline and analyzed by a laboratory-developed RT-PCR test were used to evaluate Sofia (Quidel), Standard Q (SD Biosensor), and Panbio™ (Abbott) rapid antigen detection tests (RADTs). A subset of the specimens was subjected to virus culture.

**Results:** The specificity of all RADTs was 100% and the sensitivity and percent agreement was 80% and 85% for Sofia, 81% and 85% for Standard Q, and 83% and 86% for Panbio™, respectively. All three RADTs evaluated in this study reached a more than 90% sensitivity for samples with a high viral load as estimated from the low Ct values in the reference RT-PCR. Virus culture was successful in 80% of specimens with a Ct value <25.

**Conclusions:** As expected, the RADTs were less sensitive than RT-PCR. However, they benefit from the speed and ease of testing, and lower price as compared to RT-PCR. Repeated testing in appropriate settings may improve the overall performance.

## Introduction

The superior analytical sensitivity and specificity makes RT-PCR the primary diagnostic tool for the detection of SARS-CoV-2 in respiratory samples [1-2]. Rapid antigen detection tests (RADTs) can be cheap and easy to use compared to RT-PCR. Contrary to PCR, antigen tests do not amplify the detected target, rendering them generally less sensitive [3]. The SARS-CoV-2 RADTs are relatively new in the market, without extensive evaluation. They appear to show very variable performance: according to a systematic review published in August 2020, RADT sensitivity values varied from 0% to 94% [4]. We evaluated three CE IVD marked SARS-CoV-2 RADTs: Sofia (Quidel), Standard Q COVID-19 Ag (SD Biosensor), and Panbio™ (Abbott). In each of these tests, the sample is suspended or diluted in a sample inactivation medium and transferred to a test device. If SARS-CoV-2 antigen is present above a threshold concentration, a line will appear in the test device within 15 minutes. For Panbio™ and Standard Q, the line is visible for a naked eye, and for Sofia, detection of the fluorescent signal is automated.

## Materials and methods

The evaluation was performed at the HUS Diagnostic Center, the diagnostic laboratory of Helsinki University Hospital and Helsinki and Uusimaa Hospital District, Helsinki, Finland. Research permit HUS/157/2020 (Helsinki University Hospital, Finland) was obtained from the local review board.

### Evaluated tests

We evaluated the performance of Quidel Sofia SARS FIA (Quidel, San Diego, CA), Standard Q COVID-19 Ag test (SD Biosensor, Republic of Korea), and Panbio™ (Abbott Diagnostic GmbH, Jena, Germany) according to each manufacturer’s guidelines for samples in virus transport medium. All evaluated tests are intended for fresh swab samples, so this is off-label use and leads to dilution of samples, see Table 1.

**Table 1.** Main properties of the three RADTs evaluated in this study.

|  | **Quidel Sofia SARS Antigen FIA** | **SD Biosensor SARS-CoV-2 Rapid Antigen test** | **Abbott Panbio™ COVID-19 Ag Rapid TEST Device NP** |
| --- | --- | --- | --- |
| **Intended use** | Nasopharyngeal and nasal swabs | Nasopharyngeal swabs | Nasopharyngeal swabs |
| **Target** | Nucleocapsid protein | Nucleocapsid protein | Nucleocapsid protein |
| **Interpretation of results** | Sofia or Sofia2 instrument | Visual | Visual |
| **Procedural control/visible control line** | Yes/No | Yes/Yes | Yes/Yes |
| **External controls** | Yes | Available, but not included in the kit | Yes |
| **LoD, as reported by the manufacturer** | $1.13 \times 10^2$ TCID<sub>50</sub>/mL | $3.12 \times 10^{2.2}$ TCID<sub>50</sub>/mL | $2.5 \times 10^{1.8}$ TCID<sub>50</sub>/mL |
| **Sensitivity, as reported by the manufacturer** | 96.67% | 96.52% | 93.33% |
| **Specificity, as reported by the manufacturer** | 100 % | 99.68% | 99.45% |
| **Incubation time** | 15 minutes | 15-30 minutes | 15-20 minutes |
| **Sample volume** | 120 µl | 3 drops | 5 drops |
| **Sample volume when using a frozen VTM sample µl** | 250 | 350 | 150 |
| **Sample dilution when using a frozen saline sample in this study, approximately** | 1:6 | 1:4 | 1:10 |
| **Regulatory certification** | CE-IVD, FDA EUA | CE-IVD | CE-IVD |
| **Lots used in this evaluation** | 143489 | QCO3020105 | 41ADF024A |

### Reference test

The samples were originally analyzed with a laboratory-developed RT-PCR test (LDT) based on the method by Corman and others [5] and modified by us [6] to detect the N gene target of SARS-CoV-2.

### Patient samples

A total of 198 nasopharyngeal swabs in 0.9% saline taken between April and November 2020 and stored in −20°C were available for the study.

The analytical performance evaluation was conducted with 102 samples: 40 LDT negative and 62 positive samples, selected to cover a wide Ct value range. The Ct value of the positive control varied between 26.35-32.66.

To investigate the test performance in an outpatient setting, another set of LDT positive samples from adults from outpatient clinics and drive-through testing sites was included by selecting samples systematically backwards from 18 November until 96 specimens were reached, i.e. November 1-18 2020. Samples from hospitals and occupational health services were excluded, as well as samples that had less than the required 750 µl for three RADTs left. The positive control Ct values varied from 25.56 to 27.96. Eighty-seven samples were analyzed by Sofia, 90 by Panbio™ and 96 by Standard Q.

### Virus culture

The PCR positive subset of samples used for analytical performance evaluation was subjected to virus isolation experiments in Vero E6 TMRPSS2 cells as described in [7].

### Statistics

Concordance of the results was examined in McNemars’s test. Level of statistical significance was set at P<0.05. Cohen’s kappa coefficient was computed to assess the agreement between the methods by chance and Mann-Whitney U test was used to compare the Ct value medians. Statistical analysis was performed using SPSS/PASW statistical program package, version 25 (IBM SPSS Inc., Chicago, IL, USA).

## Results

The performance of the Sofia, Standard Q and Panbio™ RADTs in comparison with the LDT RT-PCR was assessed by analysing 188, 198 and 190 specimens, respectively. The three RADTs provided valid results for all specimens. The specificity for all three tests was 100%: all RT-PCR negative samples tested negative. In all, the results of Sofia were 80.4% (119/148) consistent with the reference RT-PCR with a kappa value of 0.636 (P<0.001). The Standard Q and Panbio™ tests achieved overall agreements of 84.85% and 86.32% as compared to the RT-PCR with kappa values of 0.633 (P<0.001) and 0.660 (P<0.001), respectively. For all of the RADTs, the observed difference to the reference PCR was statistically significant (P<0.001). The median Ct values of the specimens positive with RADTs were significantly lower than those of the specimens with false negative results (P<0.001 for all RADTs) (Table 2 and Figure 1).

**Table 2.** The performance of RADTs, all samples, all RT-PCR positive samples, and samples grouped according to Ct values of the N gene target in the LDT RT-PCR. Tested, n, number of tested samples. Same as ref: number of samples tested the same as reference LDT RT-PCR test, positive or negative. Tested pos, n, number of samples that were positive in RADT. Ag^-^/Ag^+^: Ct median of samples that

|  |  | SOFIA<br>QUIDEL | STANDARD Q<br>SD BIOSENSOR | PANBIO<br>ABBOTT |
| --- | --- | --- | --- | --- |
| All samples | Tested, n | 188 | 198 | 190 |
|  | Same as ref | 159 | 168 | 164 |
|  | Agreement | 84.57% | 84.85% | 86.32% |
|  | 95% CI | 78.72-89.04 | 79.19-89.18 | 80.70-90.49 |
| All RT-PCR positive samples | Tested, n | 148 | 158 | 152 |
|  | Tested pos, n | 119 | 128 | 126 |
|  | Sensitivity | 80% | 81% | 83% |
|  | Agreement | 80.41% | 81.01% | 82.89% |
|  | 95% CI | 73.28-86.00 | 74.19-86.36 | 76.12-88.05 |
|  | Ct median (Ag<sup>-</sup>/Ag<sup>+</sup>) | 31.75/20.97 | 31.17/20.95 | 31.08/20.98 |
| RT-PCR positives, Ct <25 | Tested, n | 89 | 97 | 92 |
|  | Tested pos, n | 88 | 96 | 90 |
|  | Sensitivity | 99% | 99% | 98% |
| RT-PCR positives, Ct 25-29.99 | Tested, n | 34 | 35 | 34 |
|  | Tested pos, n | 28 | 24 | 26 |
|  | Sensitivity | 82% | 69% | 76% |
| RT-PCR positives, Ct <30 | Tested, n | 123 | 132 | 126 |
|  | Tested pos, n | 116 | 120 | 116 |
|  | Sensitivity | 94% | 91% | 92% |
| RT-PCR positives, Ct ≥30 | Tested, n | 25 | 26 | 26 |
|  | Tested pos, n | 3 | 8 | 10 |
|  | Sensitivity | 12% | 31% | 38% |

**Figure 1.**
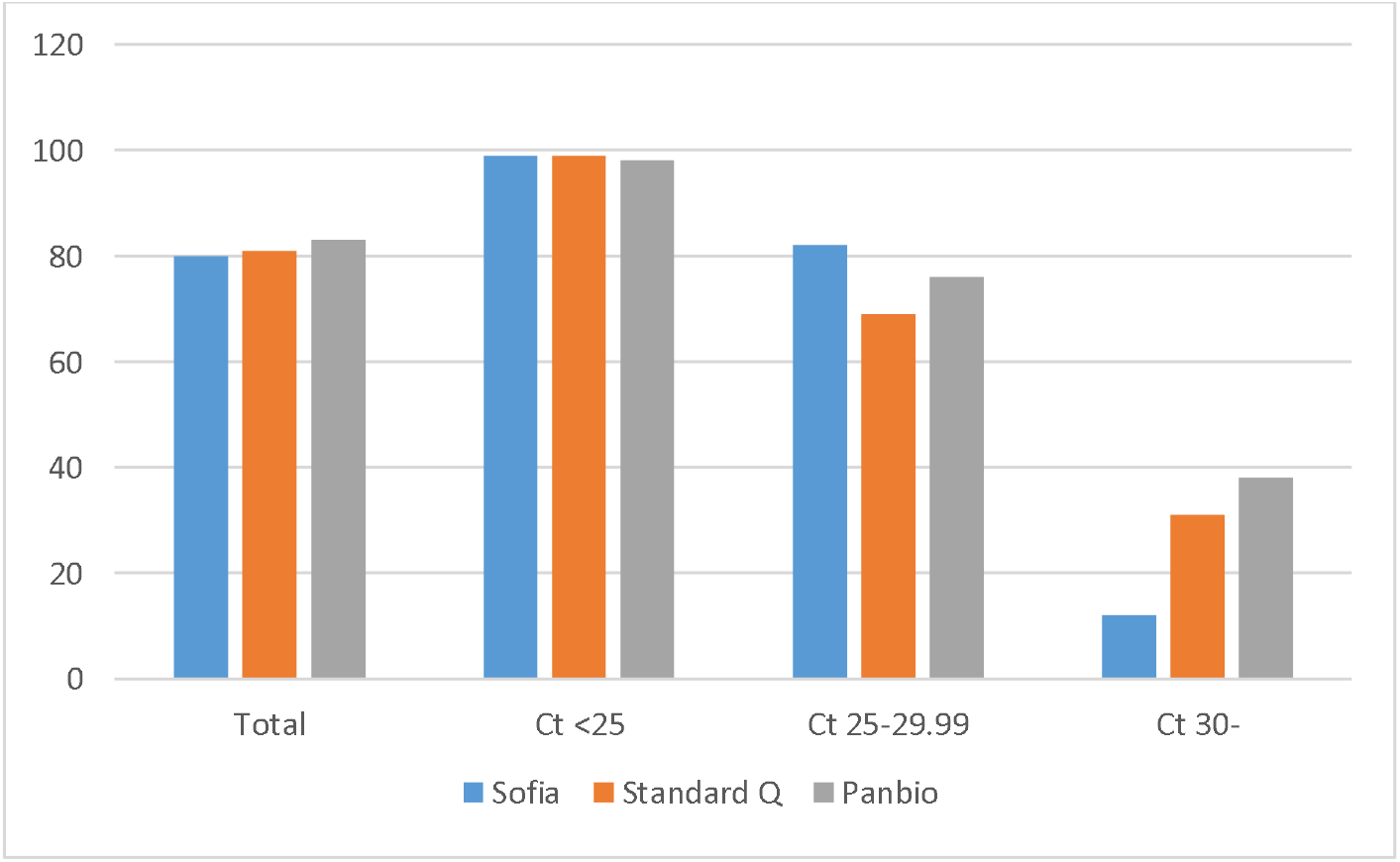
Sensitivities of Sofia, Standard Q and Panbio rapid antigen detection tests for SARS-CoV-2. Total number of RT-PCR positive samples tested for Sofia: 148, Standard Q: 158, Panbio: 152; number of RT-PCR positive samples, Ct<25, tested for Sofia: 89, Standard Q: 97, Panbio: 92. Number of RT-PCR positive samples, Ct 25-29.99, tested for Sofia: 34, Standard Q: 35, Panbio: 34. Number of R-PCR positive samples, Ct ≥30, tested for Sofia: 25, Standard Q: 26, Panbio: 26.

Table 3 shows the results for the analytical evaluation of the tests, 62 positive and 40 negative samples in LDT RT-PCR with results of virus culture experiments. Of 18 culture positive specimens, 17 were positive in RADTs.

**Table 3.** The performance of the antigen tests and virus culture in a panel of selected 62 positive and 40 negative samples according to Ct values of the N gene target in the LDT RT-PCR. Tested, n, number of tested samples. Tested pos, n, number of samples that were positive in antigen test or virus culture. ND, not done.

|  |  | SOFIA<br>QUIDEL | STANDARD Q<br>SD BIOSENSOR | PANBIO<br>ABBOTT | VIRUS<br>CULTURE |
| --- | --- | --- | --- | --- | --- |
| Total RT-PCR positive samples | Tested, n | 61 | 62 | 62 | 59 |
|  | Tested pos, n | 39 | 43 | 48 | 18 |
|  | Sensitivity | 64% | 69% | 77% | 31% |
| RT-PCR positives, Ct <25 | Tested, n | 22 | 22 | 22 | 20 |
|  | Tested pos, n | 22 | 22 | 22 | 16 |
|  | Sensitivity | 100% | 100% | 100% | 80% |
| RT-PCR positives, Ct 25-29.99 | Tested, n | 19 | 19 | 19 | 19 |
|  | Tested pos, n | 15 | 14 | 16 | 2 |
|  | Sensitivity | 79% | 74% | 84% | 11% |
| RT-PCR positives, Ct <30 | Tested, n | 41 | 41 | 41 | 39 |
|  | Tested pos, n | 37 | 36 | 38 | 18 |
|  | Sensitivity | 90% | 88% | 93% | 46% |
| RT-PCR positives, Ct ≥30 | Tested, n | 20 | 21 | 21 | 20 |
|  | Tested pos, n | 2 | 7 | 10 | 0 |
|  | Sensitivity | 10% | 33% | 48% | 0% |
| RT-PCR negatives | Tested, n | 40 | 40 | 38 | ND |
|  | Tested pos, n | 0 | 0 | 0 |  |

The Ct values in the 96 positive samples of the adult outpatients were 10.74-32.49. In all, 75 samples (78%) had a Ct value <25, sixteen (17%) had Ct values of 25-29.99 and five (5%) ≥30. The sensitivity values of RADTs were 87% (Panbio™), 89% (Standard Q) and 92% (Sofia) but reached 92-96% for samples with Ct <30. (Table 4 and Figure 2).

**Table 4.** The performance of antigen tests in a panel of positive adult outpatient samples in November 1-18, 2020, grouped according to Ct values of the N gene target in the LDT RT-PCR. Tested, n, number of tested samples. Tested pos, n, number of samples that were positive in antigen test.

|  |  | SOFIA<br>QUIDEL | STANDARD Q<br>SD BIOSENSOR | PANBIO<br>ABBOTT |
| --- | --- | --- | --- | --- |
| Total RT-PCR positive samples | Tested, n | 87 | 96 | 90 |
|  | Tested pos, n | 80 | 85 | 78 |
|  | Sensitivity | 92% | 89% | 87% |
| RT-PCR positives, Ct <25 | Tested, n | 67 | 75 | 70 |
|  | Tested pos, n | 66 | 74 | 68 |
|  | Sensitivity | 99% | 99% | 97% |
| RT-PCR positives, Ct 25-29.99 | Tested, n | 15 | 16 | 15 |
|  | Tested pos, n | 13 | 10 | 10 |
|  | Sensitivity | 87% | 63% | 67% |
| RT-PCR positives, Ct <30 | Tested, n | 82 | 91 | 85 |
|  | Tested pos, n | 79 | 84 | 78 |
|  | Sensitivity | 96% | 92% | 92% |
| RT-PCR positives, Ct ≥30 | Tested, n | 5 | 5 | 5 |
|  | Tested pos, n | 1 | 1 | 0 |
|  | Sensitivity | 20% | 20% | 0% |

**Figure 2.**
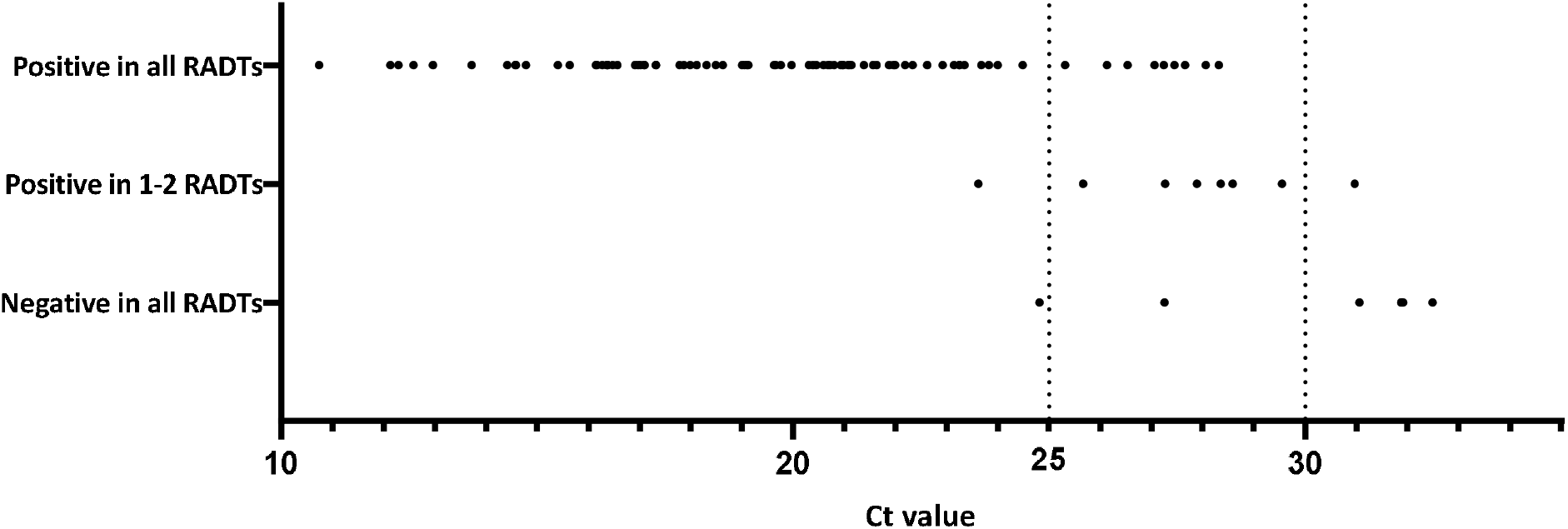
Sensitivity of the RADTs in comparison to Ct values in RT-PCR. Samples include all the LDT RT-PCR positive nasopharyngeal samples from adult outpatients at HUSLAB in 1.-18. November 2020 that had enough sample volume left for three antigen tests. N= 96, however, in some cases, not all three RADTs were performed to all samples. Each dot indicates one samples. Ct <25: 75 samples; Ct 25-29.99: 16 samples; Ct ≥30: 5 samples.

## Discussion

We evaluated three lateral-flow RADTs for the detection of SARS-CoV-2 infection in nasopharyngeal swab samples collected in 0.9% saline. Our results agree with previous observations that RADTs perform well in detecting culture positive samples and samples with high viral loads [8-11]. Compared to RT-PCR, the sensitivity of RADTs is low: in the present study the sensitivity was only 12-38% in samples with Ct≥30 (Table 2).

However, the distribution of Ct values of RT-PCR positive samples in settings where antigen detection tests could primarily be used is unknown. We wanted to approach the real-life setting by testing all positive outpatient adult samples tested by LDT in our laboratory during 2.5 weeks in November 2020. The testing strategy in Finland in November 2020 assumed patients to have at least mild symptoms of SARS-CoV-2 infection. Thus, this panel is unlikely to include asymptomatic or presymptomatic patients. During the time of evaluation, the sampling was available in average within 12 hours of seeking for testing. Although medical records of outpatients were not available, we assume the majority of this panel to consist of samples taken within a couple of days from symptom onset. 78% of samples had a Ct value <25, and 95% <30, of the N gene in our LDT. This may justify using antigen tests as a diagnostic tool even when their sensitivity compared to nucleic acid testing is poor for samples with high Ct values. Careful consideration between the advantages (ease, speed, costs) and disadvantages (lower sensitivity) needs to be conducted in the selection of approriate testing strategies [11-12]. If RADTs are considered, laboratories need to perform independent clinical validations, as manufacturers’ declaration of performance may not include satisfactory selection of clinical samples. WHO has stated SARS-CoV-2 a biohazard risk for laboratory personnel [13], which needs to be accounted for in RADT testing. In addition, RATDs are not easily scalable for mass testing.

The present study is limited by its retrospective nature. Ideally, samples for RADT evaluation should be collected according to manufacturer’s instructions. Instead, we used frozen samples in saline. This may lead to underestimation of the sensitivity of the antigen detection tests.

All three RADTs, Sofia, Standard Q and Panbio™, were specific but less sensitive than RT-PCR. In appropriate settings, this disadvantage may be compensated by the ability to perform the test outside a central laboratory, the shorter turnaround time, and lower price. The sensitivity of the testing regimen as a whole can be increased by repeat testing [14]. Simple and cheap RADTs with a satisfactory performance, such as the three tests evaluated in this study, make repeated testing a realistic diagnostic approach in settings in which RT-PCR is not readily or quickly available.

## Data Availability

All detailed data is available from the authors upon request.

## Acknowledgements

We thank Marianne Ahola, Jenna Roivas and Laila Shakari for excellent technical assistance.

